# Development and Evaluation of *iSupport-Malaysia*: A Multimedia Web-Based Psychoeducational Intervention for Dementia Caregivers

**DOI:** 10.64898/2026.04.14.26350743

**Authors:** Ken Joey Loh, Wan Ling Lee, Alvin Lai Oon Ng, Felicia Fei Lei Chung, Elil Renganathan

**Affiliations:** Faculty of Medical and Life Sciences, Sunway University, Sunway City, 47500 Petaling Jaya, Selangor, Malaysia; Faculty of Medicine, Universiti Malaya, 50603 Kuala Lumpur, Malaysia; Jeffrey Cheah School of Medicine and Health Sciences, Monash University Malaysia, 47500 Petaling Jaya, Selangor, Malaysia

**Author notes:** **Corresponding Author**: Alvin Lai Oon Ng, DPsych, School of Psychology, Faculty of Medical and Life Sciences, Sunway University, Sunway City, Malaysia. **Note:** This manuscript is currently submitted and under consideration at *Informatics for Health and Social Care*.

**Keywords:** caregivers, dementia, usability, digital health intervention, psychoeducation, cultural adaptation

## Abstract

**Background:** Caring for people with dementia can impose a considerable psychological burden on caregivers, yet access to caregiver support in Malaysia remains limited. The World Health Organization’s *iSupport for Dementia* program provides dementia education via textual, e-learning format. However, a culturally adapted Malaysian version has not been available.

**Objective:** This study aimed to develop and gather user feedback on a culturally adapted, multimedia version of iSupport tailored for Malaysia (iSupport-Malaysia).

**Methods:** Guided by a four-phase cultural adaptation framework, the generic iSupport content was translated into Bahasa Malaysia, adapted to local customs, and transformed into multimedia lessons on an e-learning platform. A mixed-methods design was used to explore user perceptions and evaluate usability through four homogeneous focus group discussions and 15 individual usability test sessions with informal caregivers (FG: *n*=9; UT: *n*=9) and healthcare professionals (FG: *n*=11; UT: *n*=6). Focus groups examined aesthetics, ease of use, clarity, cultural relevance, comprehensiveness, and satisfaction. Usability testing involved Think Aloud tasks, post-test questionnaires, and brief interviews. Qualitative data was analysed thematically, and descriptive statistics summarised usability performance.

**Results:** iSupport-Malaysia demonstrated good usability (*M*=74.3±18.0), with most tasks completed without assistance. Strengths included interactive learning activities, peer discussion features, and flexible self-paced learning. Content was viewed as culturally appropriate, credible, and useful. Suggested improvements included enhancing visual aesthetics, shortening videos, refining quizzes, and increasing practical relevance.

**Conclusion:** User insights indicate that iSupport-Malaysia is usable and culturally appropriate. These findings will inform refinement of the platform prior to the pilot feasibility study and provide recommendations for future multimedia-based caregiver interventions.

## 1. Introduction

Dementia is a neurocognitive disorder that progressively causes severe impairments in memory, thinking, and daily functioning. Globally, over 57 million people were diagnosed with dementia in 2021; among them, 60% resided in lower- and middle-income countries (World Health Organization, 2024). From an economic perspective, dementia costs are expected to reach USD 2.8 trillion globally by 2030, with half of this attributed to informal caregiving (Alzheimer Disease International, 2025).

In Malaysia, the rising ageing population is expected to bring about 249% increase in dementia cases from 142,172 in 2019 to 411,045–596,328 by 2050 (Nichols et al., 2022). Due to the filial piety culture in Southeast Asian countries, informal caregiving of people with dementia is often carried out by family members at home. A recent cross-sectional study conducted in 6 tertiary hospitals in Malaysia reported that informal caregivers of people living with Alzheimer’s disease spent an average of 3.85–11.77 hours per day providing care, including assistance with basic and instrumental activities of daily living and supervision (Ong et al., 2025). Coupled with the inadequate knowledge of behavioural-psychological symptoms management and lack of access to dementia care services and resources (including respite care and education), informal caregivers are especially vulnerable to high caregiving stress, burden, social isolation, and reduced quality of life (Bui et al., 2022; Loh et al., 2024; Mohd Razi et al., 2023).

Advancements in information and communication technologies (ICT) have positioned digital education, particularly e-learning platforms, as a convenient and useful tool for delivering health education across geographical barriers. These platforms enable learners to access information in a flexible and engaging way using modern digital devices. Unlike conventional face-to-face training or classroom-based approaches, e-learning utilises modular and interactive features through a digital format to enhance learning processes, thereby potentially improving knowledge retention (Mugil et al., 2025). Additionally, it can overcome barriers such as costs and transportation for attending in-person sessions, and inability to leave home because of caregiving roles, which in turn improves uptake (Gonzalez-Fraile et al., 2021).

The iSupport for Dementia program, developed by the World Health Organization (WHO), is a digital intervention that delivers evidence-based dementia educational content via a web-based, e-learning platform. With twenty three lessons categorised under five modules (see Figure 1), this self-paced program aims to equip caregivers with practical strategies to manage the behavioural/ psychological symptoms of dementia and activities of daily living, enhance emotional self-management, and improve communication skills (World Health Organization, 2019). A distinctive feature of this program is its self-paced format, which could be beneficial for informal caregivers who are required to balance multiple roles and commitments. Nonetheless, the generic iSupport does not include peer interaction, and all 23 lessons are delivered in a heavily text-based format rather than using interactive or multimedia elements. Although this instructional design makes iSupport scalable and easy to translate, several iSupport adaptation studies have highlighted limitations in sustaining learners’ engagement over time, citing that more engaging multimedia formats, such as audios or videos, could be more helpful in increasing motivation to use (Baruah et al., 2020; Molinari-Ulate et al., 2023; Xiao et al., 2022).

**Figure 1.**
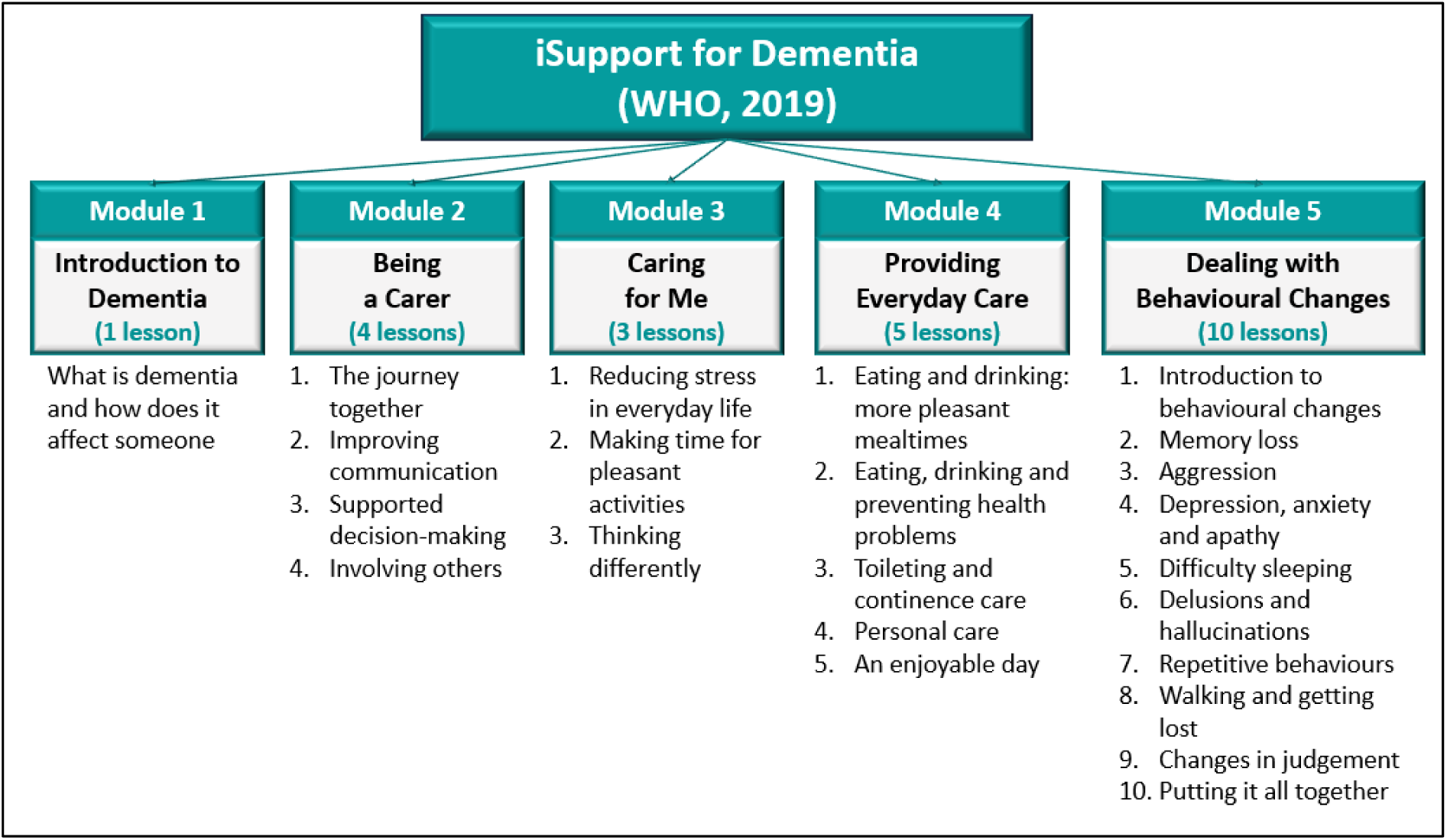
Overview of the 23 lessons (parked under five modules) in the generic iSupport. Diagram adapted from WHO’s iSupport for Dementia training manual (World Health Organization, 2019).

Since its inception in 2019, iSupport has been culturally adapted for use in 57 countries and translated into 37 languages (World Health Organization, 2024). Cultural adaptation refers to “the systematic modification of an evidence-based treatment or intervention protocol to consider language, culture, and context in such a way that it is compatible with the client’s cultural patterns, meanings, and values” (p. 362)(Bernal et al., 2009). In dementia care, caregiving practices are shaped not only by knowledge of ageing and cognitive impairment, but also by cultural expectations of filial duty and family dynamics. Therefore, to increase acceptance and engagement among local dementia caregivers, culturally adapted interventions need to tailor the content according to the literacy levels, language, local customs and caregiving practices (Ibekaku et al., 2025). The adaptation process shall also incorporate the perspectives of both end-users (i.e. caregivers) and local professionals through iterative feedback and testing (Teles et al., 2021).

### 1.1 Aim & Objectives

In line with the Global Action Plan on the Public Health Response to Dementia 2017–2025, the World Health Organization recommends that every country provide at least one accessible, evidence-based support and training programme for informal dementia caregivers (World Health Organization, 2017). Malaysia has taken steps in this direction through the National Dementia Action Plan 2023–2030 (Ministry of Health Malaysia, 2024), which prioritizes strengthening caregiver and care-partner support. Yet, accessible and scalable psychoeducational resources for Malaysian caregivers remain scarce. To the best of authors’ knowledge, only two local initiatives have been documented to-date. The Demensia-KITA mobile application offers culturally tailored dementia information and service directories in national Malay language (Mahfar et al., 2025; Rashid et al., 2024), but its content is largely text-based, minimally interactive, and its usability and effectiveness have yet to be evaluated (Mahfar et al., 2025; Rashid et al., 2022). Another study delivering iSupport content via telephone showed reductions in caregiver burden, anxiety and distress (Ahmad et al., 2024). However, this approach is labour-intensive as it was dependent on trained healthcare personnels, making it difficult to scale beyond clinical settings. These limitations collectively highlight the need for an engaging, accessible eHealth intervention that can reach caregivers across regions and literacy levels without relying on specialist manpower.

The present study aimed to develop an iSupport program tailored to the Malaysian cultural context, hereafter referred to as iSupport-Malaysia. Using a mixed methods approach, the cultural adaptation process of the generic iSupport program was guided by Barrera and Casrtro’s (2006) four-phase cultural adaptation framework (Barrera & Castro, 2006) and the iSupport Adaptation and Implementation Guidelines (World Health Organization, 2018). The objectives were: (i) to explore users’ perceptions of the preliminary iSupport-Malaysia e-learning platform, particularly regarding its usability and overall digital learning experience; and (ii) gather users’ feedback on the perceived appropriateness of the adapted lesson content. Insights gathered from this study will help identify strengths, gaps, and recommendations to inform further refinement prior to implementation in our subsequent pilot feasibility clinical trial.

## 2. Methods

Figure 2 outlines the research phases of this study, adapted from Barrera and Castro’s framework (2006) on cultural adaptation (Barrera & Castro, 2006). The essential elements of the adaptation model are: (i) local translation; (ii) preliminary adaptation design; (iii) preliminary adaptation tests; and (iv) adaptation refinement.

**Figure 2.**
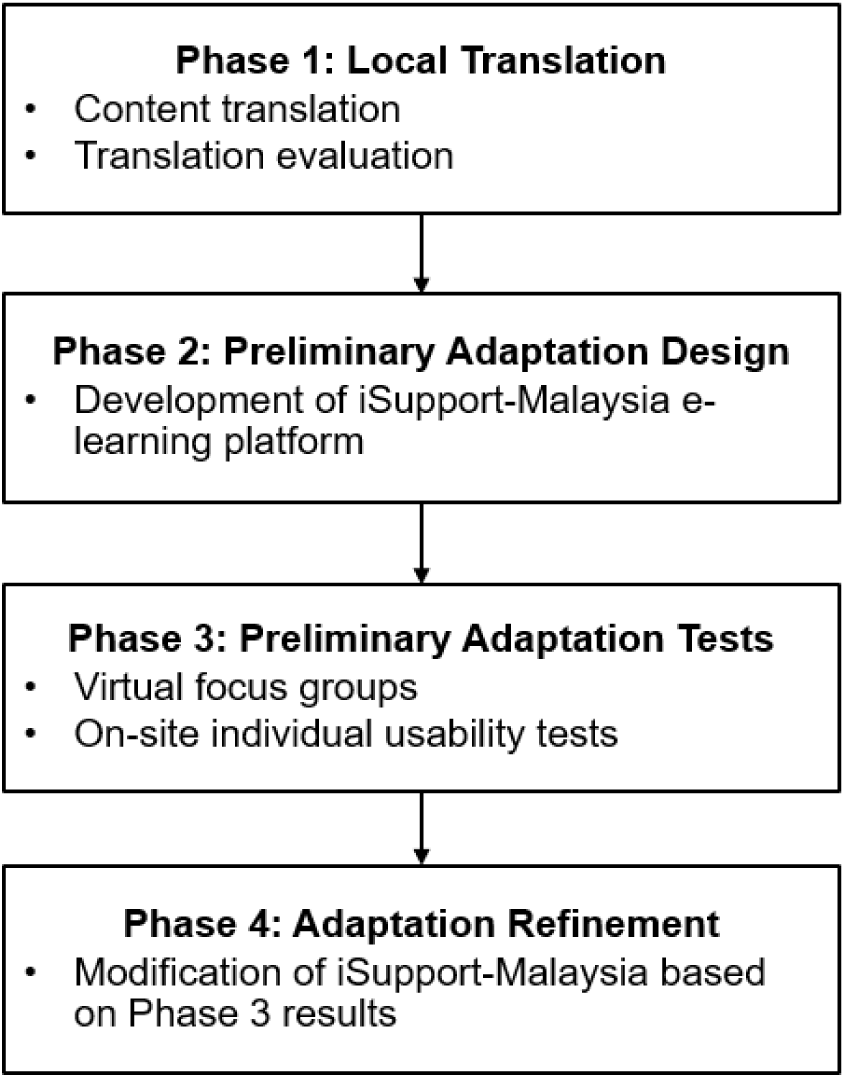
Flow chart outlining the four phases of cultural adaptation, adapted from Barrera and Castro’s (2006) framework.

### 2.1 Phase 1: Local Translation

The first phase in our adaptation framework was renamed as ‘local translation’ instead of ‘information gathering’, as our focus for this phase was to translate the generic iSupport program content from English to Bahasa Malaysia (i.e. the national language in Malaysia). Initial translation of the generic iSupport was conducted by five bilingual nurses using forward translation (from source language to local language). Following that, the translations were reviewed by two bilingual clinical experts in dementia care. Subsequently, the translated materials were appraised for translation accuracy and refined for translation relevance, via discussions with stakeholders’ group comprising four geriatricians, three nurses, two rehabilitation professionals, one gerontologist and two informal caregivers.

### 2.2 Phase 2: Preliminary Adaptation Design

In this phase, the iSupport Adaptation and Implementation Guidelines (World Health Organization, 2018) were used to guide the development of the e-learning materials for the translated iSupport modules. To ensure conceptual and content fidelity with the generic version while enhancing cultural relevance for Malaysian caregivers, linguistic and contextual adaptations were made by the first author (KJL), including the modification of words and expressions (e.g., food, household items, and attire) to reflect local usage. Character names in the case scenarios were localised to represent Malaysia’s three major ethnic groups (Malay, Chinese, and Indian), and caregiving scenarios were revised to emphasise family-based care, since it is a common practice in Malaysia for family members to take care of older adults at home, with or without the assistance of home helpers and paid caregivers (Goodson et al., 2021). A comprehensive list of local resources, such as memory clinics, dementia associations, and non-profit daycare centres, was also added. Additionally, the lesson language was simplified to improve comprehensibility. These revisions were reviewed by a senior investigator experienced in dementia care (WLL) and a representative from a local dementia organization, and final changes were made upon consensus.

Previous iSupport adaptation work indicated that caregivers preferred lesson formats that are more engaging, such as audio and video presentations (Baruah et al., 2020; Molinari-Ulate et al., 2023; Xiao et al., 2022). Therefore, we transformed all the 23 lessons into multimedia formats, designed to be engaging and bite-sized to facilitate learning. Video scripts for all 23 lessons were developed and refined collaboratively within the research team before production by an animation video creator. Each animation video conveyed the lesson’s key concepts within a duration of five to nine minutes. Subsequently, the completed videos were hosted on the OpenLearning^TM^ e-learning platform. While retaining the original five-module structure of iSupport, each lesson was presented in the following sequential components: learning objectives, animation video and interactive quiz with immediate feedback. For certain lessons, open-ended reflection exercises (peer sharing or self-administered worksheet format) and downloadable handouts (infographics or reading materials) were also included. To encourage peer learning, the ‘comment’ features were included in all lessons for users to exchange views and knowledge. Users will also receive a downloadable certificate upon completing all 23 lessons. Essentially, our instructional design for iSupport-Malaysia follows established e-learning pedagogical principles of behaviourism, cognitivism and constructivism, where learning is reinforced and made engaging through features like multimedia educational materials, assessments (quizzes) and collaborative peer discussion to enhance self-directed learning (Jabsheh, 2024; Mugil et al., 2025).

### 2.3 Phase 3: Preliminary Adaptation Tests

This phase involved pilot testing the preliminary version of iSupport-Malaysia e-learning platform. Data collection was carried out in two parts, using a **mixed methods** approach, to address two research questions: (i) How do users perceive their digital learning experience with iSupport-Malaysia? and (ii) What are users’ views on the educational content in iSupport-Malaysia? Qualitative reporting of this phase followed the Consolidated Criteria for Reporting Qualitative Studies (COREQ) guideline (Tong et al., 2007) (<u>Supplemental File</u> <u>1: COREQ checklist)</u>.

Adapted from the iSupport-Portugal usability study protocols (Teles et al., 2021), there are two parts to the methodology. The first part involved virtual, homogeneous focus group discussions with caregivers and healthcare professionals separately, where we obtained in-depth user feedback regarding the digital learning experience and content appropriateness. Homogeneity was maintained within groups as this could encourage more open sharing among participants who perceived having similar experiences and backgrounds with one another (Wong, 2008). The second part comprised a series of individual usability tests on the iSupport-Malaysia platform, where we collected real-time feedback (quantitative and qualitative) when participants were navigating the platform.

#### 2.3.1 Participants

Potential caregivers who met the following inclusion criteria were recruited via purposive sampling through social media and referrals from local dementia associations: a) Malaysian adults aged ≥ 18 years; b) proficient in written Malay; c) family members or paid caregivers currently providing care to a person with dementia for at least six months; d) frequent internet users (≥ 2 days per week); and e) owned an email address. On the other hand, healthcare professionals were recruited through purposive sampling from researchers’ professional networks. Eligibility criteria were: a) Malaysian adults aged ≥ 18 years; b) professionally trained in any of these disciplines (geriatric and gerontology, medicine, nursing, psychology, social work); and c) had ≥ 1 year of experience supporting people with dementia and their families. For the on-site usability tests, both caregivers and healthcare professionals must reside in Selangor and Kuala Lumpur regions. Nine caregivers^a^ and eleven healthcare professionals provided electronic informed consent for the virtual focus groups, while another nine caregivers and six healthcare professionals provided written informed consent for the individual usability tests. Their demographic characteristics were summarised in Table 1 under <u>Results</u>.

**Table 1.**
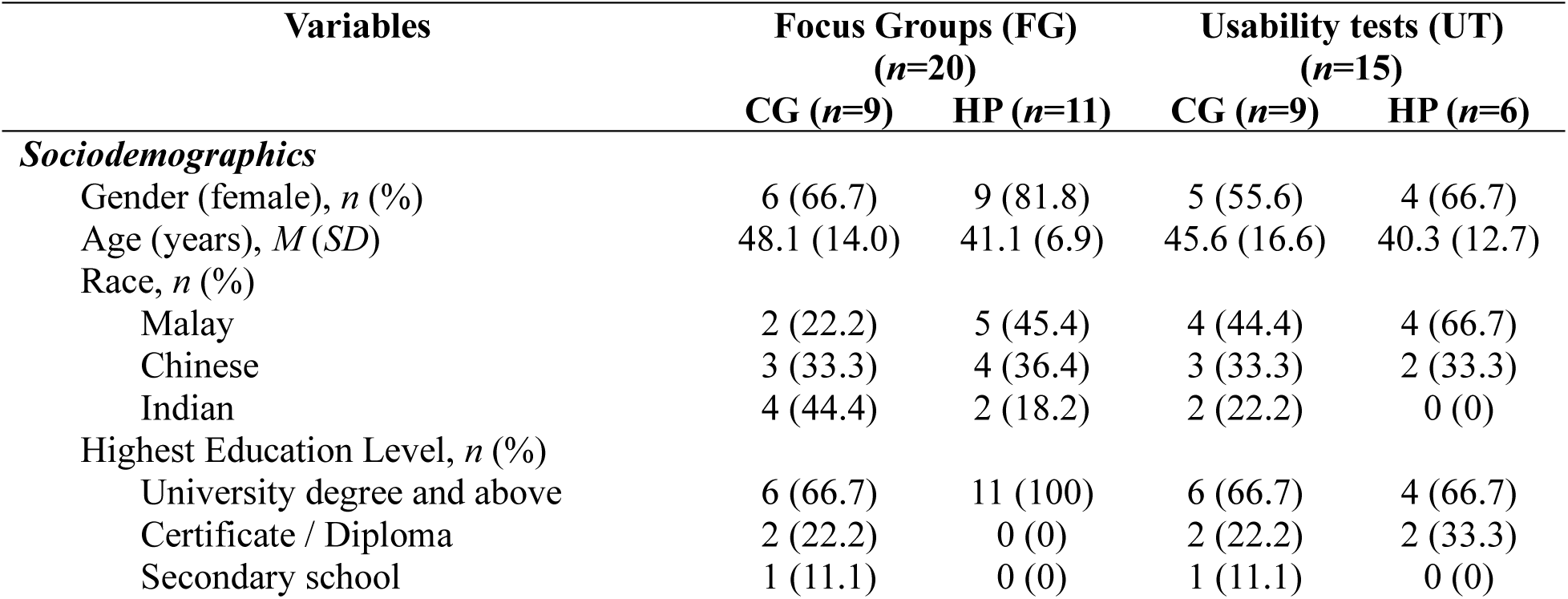
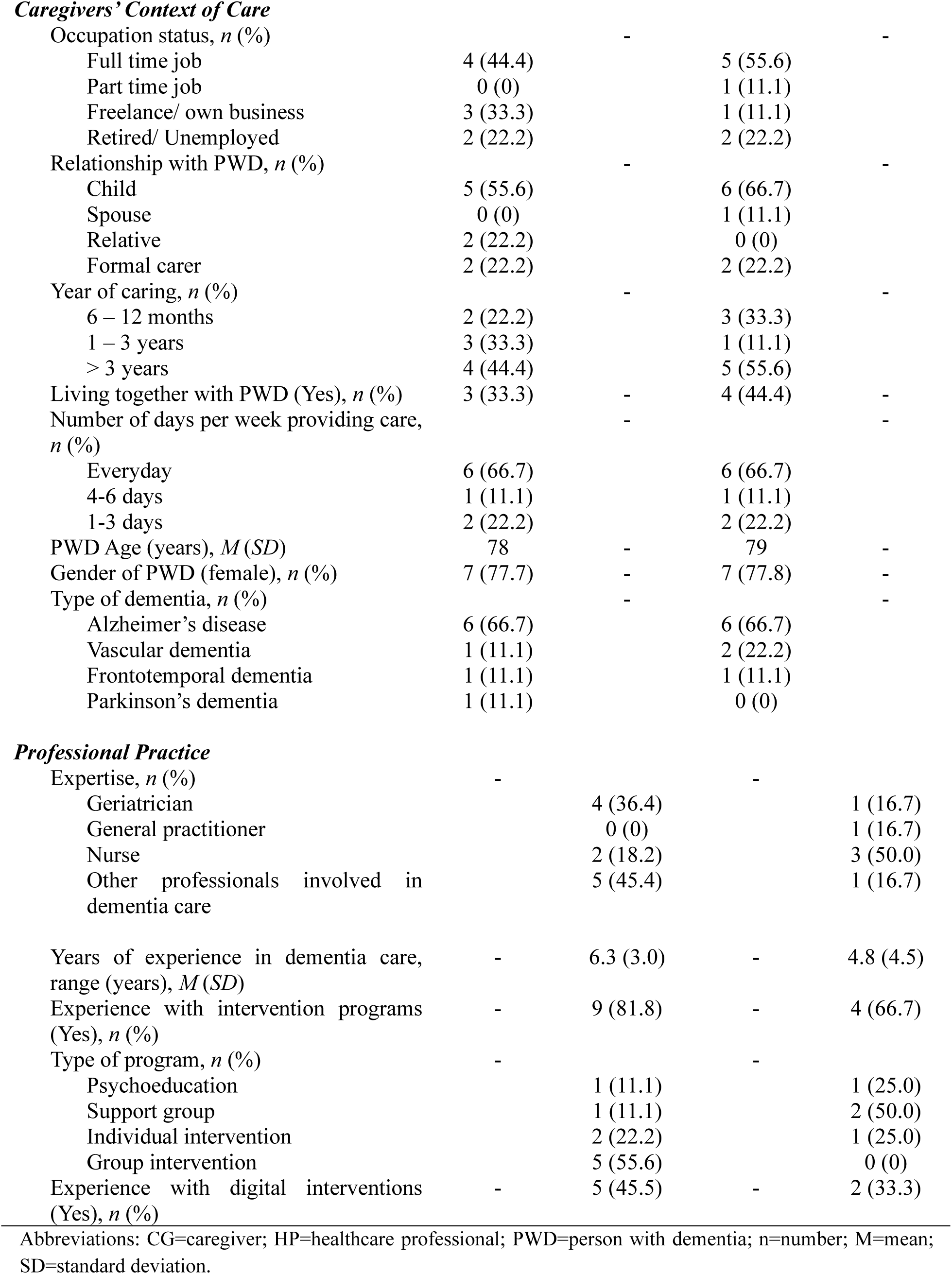
Socio-demographic characteristics of participants from preliminary adaptation tests.

| Variables | Focus Groups (FG)<br>($n=20$) | | Usability tests (UT)<br>($n=15$) | |
| --- | --- | --- | --- | --- |
| | CG ($n=9$) | HP ($n=11$) | CG ($n=9$) | HP ($n=6$) |
| ***Sociodemographics*** |  |  |  |  |
| Gender (female), $n$ (%) | 6 (66.7) | 9 (81.8) | 5 (55.6) | 4 (66.7) |
| Age (years), $M$ ($SD$) | 48.1 (14.0) | 41.1 (6.9) | 45.6 (16.6) | 40.3 (12.7) |
| Race, $n$ (%) | | | | |
| Malay | 2 (22.2) | 5 (45.4) | 4 (44.4) | 4 (66.7) |
| Chinese | 3 (33.3) | 4 (36.4) | 3 (33.3) | 2 (33.3) |
| Indian | 4 (44.4) | 2 (18.2) | 2 (22.2) | 0 (0) |
| Highest Education Level, $n$ (%) | | | | |
| University degree and above | 6 (66.7) | 11 (100) | 6 (66.7) | 4 (66.7) |
| Certificate / Diploma | 2 (22.2) | 0 (0) | 2 (22.2) | 2 (33.3) |
| Secondary school | 1 (11.1) | 0 (0) | 1 (11.1) | 0 (0) |
| **Caregivers' Context of Care** |  |  |  |  |
| Occupation status, *n* (%) |  | - |  | - |
| Full time job | 4 (44.4) |  | 5 (55.6) |  |
| Part time job | 0 (0) |  | 1 (11.1) |  |
| Freelance/ own business | 3 (33.3) |  | 1 (11.1) |  |
| Retired/ Unemployed | 2 (22.2) |  | 2 (22.2) |  |
| Relationship with PWD, *n* (%) |  | - |  | - |
| Child | 5 (55.6) |  | 6 (66.7) |  |
| Spouse | 0 (0) |  | 1 (11.1) |  |
| Relative | 2 (22.2) |  | 0 (0) |  |
| Formal carer | 2 (22.2) |  | 2 (22.2) |  |
| Year of caring, *n* (%) |  | - |  | - |
| 6 – 12 months | 2 (22.2) |  | 3 (33.3) |  |
| 1 – 3 years | 3 (33.3) |  | 1 (11.1) |  |
| > 3 years | 4 (44.4) |  | 5 (55.6) |  |
| Living together with PWD (Yes), *n* (%) | 3 (33.3) | - | 4 (44.4) | - |
| Number of days per week providing care, *n* (%) |  | - |  | - |
| Everyday | 6 (66.7) |  | 6 (66.7) |  |
| 4-6 days | 1 (11.1) |  | 1 (11.1) |  |
| 1-3 days | 2 (22.2) |  | 2 (22.2) |  |
| PWD Age (years), *M* (*SD*) | 78 | - | 79 | - |
| Gender of PWD (female), *n* (%) | 7 (77.7) | - | 7 (77.8) | - |
| Type of dementia, *n* (%) |  | - |  | - |
| Alzheimer's disease | 6 (66.7) |  | 6 (66.7) |  |
| Vascular dementia | 1 (11.1) |  | 2 (22.2) |  |
| Frontotemporal dementia | 1 (11.1) |  | 1 (11.1) |  |
| Parkinson's dementia | 1 (11.1) |  | 0 (0) |  |
| **Professional Practice** |  |  |  |  |
| Expertise, *n* (%) | - |  | - |  |
| Geriatrician |  | 4 (36.4) |  | 1 (16.7) |
| General practitioner |  | 0 (0) |  | 1 (16.7) |
| Nurse |  | 2 (18.2) |  | 3 (50.0) |
| Other professionals involved in dementia care |  | 5 (45.4) |  | 1 (16.7) |
| Years of experience in dementia care, range (years), *M* (*SD*) | - | 6.3 (3.0) | - | 4.8 (4.5) |
| Experience with intervention programs (Yes), *n* (%) | - | 9 (81.8) | - | 4 (66.7) |
| Type of program, *n* (%) | - |  | - |  |
| Psychoeducation |  | 1 (11.1) |  | 1 (25.0) |
| Support group |  | 1 (11.1) |  | 2 (50.0) |
| Individual intervention |  | 2 (22.2) |  | 1 (25.0) |
| Group intervention |  | 5 (55.6) |  | 0 (0) |
| Experience with digital interventions (Yes), *n* (%) | - | 5 (45.5) | - | 2 (33.3) |

Abbreviations: CG=caregiver; HP=healthcare professional; PWD=person with dementia; n=number; M=mean; SD=standard deviation.

#### 2.3.2 Procedure for virtual focus group discussions

Four focus group discussions (two with caregivers, two with healthcare professionals) were conducted virtually via Zoom between July and August 2024. All sessions were video-and audio-recorded.

Two weeks before the focus group sessions, participants were briefed online about the study, assisted in registering on the iSupport-Malaysia platform, and given access to assigned modules for review. To ensure adequate coverage yet minimise participants’ burden, the first caregiver and healthcare professional groups reviewed Modules 1, 2, and 4 (10 lessons), while the second groups reviewed Modules 3 and 5 (13 lessons). Participants also completed consent forms and sociodemographic questionnaires electronically prior to the sessions.

All four focus group sessions were co-facilitated by a female primary moderator (KJL), who was a psychologist trained in qualitative methods and dementia care; and a male research assistant trained in conducting interviews. Both moderators did not have any personal relationship with the participants prior to study commencement. A mock session was conducted beforehand with guidance from a senior qualitative researcher (WLL), who was a registered nurse with extensive experience in geriatric care. A semi-structured focus group guide, developed from literature and refined by the research team, focused on five domains: (1) aesthetics, (2) ease of use, (3) language clarity and cultural relevance, (4) comprehensiveness of content, and (5) user satisfaction and recommendation. The guide combined open-ended questions with voting activities to stimulate active participation in the discussion, and additional prompt questions were included if further clarification on certain topics were required. (<u>Supplemental File 2: Focus Group Guide</u>)

Sessions lasted 71–89 minutes, conducted mainly in English with Malay language interspersed as needed. All participants received a small honorarium (MYR50) for their time.

#### 2.3.3 Procedure for on-site usability tests

Usability testing evaluates the effectiveness, efficiency, and user satisfaction of a system through observing how intended users perform typical tasks on the system (International Organization for Standardization, 2019). In our study context, we observed and recorded users’ behaviours when they were completing a series of tasks on the iSupport-Malaysia website, so that any navigation issues and design flaws could be identified. To effectively capture real-time feedback during the usability test, each session was conducted individually in a quiet room at a local university. A standard Windows^TM^ laptop was used to conduct all sessions to ensure consistency. All sessions were conducted by a female researcher (KJL). Prior to the usability tests, a pilot test of the usability protocol was conducted with a medical doctor and a digital health expert. Certain instructions were refined for clarity based on feedback received.

Upon explaining study procedures and obtaining written informed consent, participants were familiarised with the ‘think aloud’ method through a short practice task on a local news website. Specifically, ‘think aloud’ required participants to imagine themselves doing the task alone at home; verbalise all thoughts, including thoughts that might be unrelated to the task; speak continuously and spontaneously without pre-planning their responses (Ericsson & Simon, 1980).

After familiarisation, the following usability tasks were administered sequentially:

1. ***Usability tasks***: Using ‘think aloud’ method, participants attempted seven tasks on iSupport-Malaysia website while spontaneously verbalising their thoughts. The tasks included account registration, accessing lessons, watching videos, completing quizzes, downloading handouts, using ‘Search’, and checking progress. Task completion was scored as ‘not completed’, ‘completed with assistance’, or ‘completed without assistance’. Participants’ verbalisations, non-verbal reactions, and navigation behaviours were recorded via written notes, audio- and screen-recordings.
2. ***Task difficulty rating***: After each task, participants rated the perceived ease of completing the task on a 5-point scale (1= ‘very difficult’; 5= ‘very easy’).
3. ***Post-test reflection:*** Participants verbally answered three open-ended questions on what they liked and disliked about the website, and suggestions for improvement.
4. ***System Usability Scale (SUS):*** The Malay-translated SUS (Mohamad Marzuki et al., 2018) was administered, which contained 10 items evaluating the usability of a web system on 5-point Likert scales (1= ‘strongly disagree’; 5= ‘strongly agree’). The term ‘mobile app’ was replaced by ‘iSupport-Malaysia website’ on all items. The final adjusted scores ranged from 0–100, with ≥68 indicating above-average usability (Brooke, 1996).
5. ***Brief questionnaire:*** The sociodemographic questionnaire from Stage 1 was administered, with an additional item asking participants to rate their IT literacy (1= ‘not confident’; 10= ‘very confident’).

Sessions were conducted in English or Malay, depending on participants’ preference. Duration of test sessions lasted for 59–76 minutes, and all sessions were audio- and video-recorded. All participants received a small honorarium (MYR30) for their time.

#### 2.3.4 Analysis

Audio recordings from focus groups and usability tests were transcribed verbatim via TurboScribe^TM^, an online AI software. All transcripts were checked against the original recordings for accuracy. As some of the transcripts contained a mix of English and Malay languages, which is a common phenomenon in Malaysia, the bilingual researcher (KJL) analysed the transcribed data in its original English and Malay languages to preserve the original meaning. Selected codes underwent forward-backward translations to ensure accuracy.

Qualitative analysis followed an inductive approach via the six-step thematic analysis framework by Braun and Clarke (2006) (Braun & Clarke, 2006), performed using Microsoft Word and Excel. Two bilingual researchers (KJL, WLL) first familiarised themselves with the data through listening to audio recordings, reading and re-reading the original transcripts, and reviewing field notes from both focus groups and usability tests. The coding process was then initiated by the primary researcher (KJL) using open coding to capture key ideas and patterns across transcripts. This was followed by iterative rounds of pattern coding, where codes were refined, merged, and grouped. Patterns were consolidated into subthemes and subsequently synthesised into overarching themes until thematic saturation was reached. To enhance analytic trustworthiness, peer debriefing was conducted between KJL, WLL and other research team members (FFLC, ALON, ER). Discrepancies were discussed and resolved through consensus during regular meetings.

Quantitative data included usability indicators (SUS scores, task completion rates, task difficulty ratings) and demographics. These data were recorded on Microsoft Excel and subsequently imported into jamovi (version 2.6), an open-source statistical software (The jamovi project, 2025). Descriptive statistics were calculated by summarising continuous variables using means and standard deviations, and categorical variables via counts and percentages.

#### 2.3.5 Qualitative and quantitative data synthesis

This study followed the triangulation mixed methods design informed by Creswell and Clark’s (2007) framework (Creswell & Clark, 2017). Qualitative (focus group discussions) and quantitative (task difficulty ratings, completion rates, SUS scores) data were collected in parallel from both caregivers and healthcare professionals, with each method providing different but complementary insights into users’ experiences with the iSupport-Malaysia platform. The two datasets were analysed separately and subsequently brought together through a process of comparative synthesis, in which qualitative themes were examined alongside quantitative usability indicators to identify areas of convergence, divergence and expansion. This integration can produce a more comprehensive understanding of the platform’s cultural relevance, usability, and areas requiring refinement.

### 2.4 Phase 4: Adaptation Refinement

Based on synthesised findings from the focus group discussions and usability testing, targeted revisions will be made to the preliminary iSupport-Malaysia web-based program. The core components of the generic iSupport will be maintained, while modifications (in terms of aesthetics, navigation, language and content) will be finalised through team consensus and stakeholder consultation. All changes, including specific changes in text or illustrations, will be documented using the WHO iSupport Local Adaptation Templates and submitted to the WHO team for review and approval prior to local pilot feasibility testing.

### 2.5 Ethical Approvals

This study was approved by the Medical Research Ethics Committee of the University of Malaya (MRECID No.: 2023927-12912) and the Research Ethics Committee of Sunway University (REC ID: PGSUREC2023/083). All participants who participated in the focus groups and usability tests were provided with information sheets to review, and all participants provided written or electronic informed consent prior to research participation.

## 3. Results

This section presents the results from the preliminary adaptation tests. This includes the demographic characteristics of the caregivers and healthcare professionals who participated in the focus groups (FGs) and usability tests (UTs), key findings from the UTs, as well as the qualitative insights gathered from participants’ feedback in both FGs and UTs.

### 3.1 Demographics

Thirty-five individuals, comprising of 18 dementia caregivers and 17 healthcare professionals, participated in the focus groups (*n*=20) or usability tests (*n*=15) (**Table 1**). For both focus groups and usability testing, caregivers were predominantly female and came from diverse age groups. Most held a university degree and were currently employed. Care recipients were mainly females, with most being diagnosed with Alzheimer’s dementia. Over half of the caregivers were children of the care recipient. Healthcare professionals were mostly female who came from diverse professional backgrounds, including geriatrics, primary care, nursing, clinical psychology, occupational therapy, physiological therapy, and care home management. Over half of the healthcare professionals had experience in developing, implementing, or evaluating intervention programs; while seven of them reported having experiences with developing, implementing or evaluating digital interventions.

All caregivers and healthcare professionals in the focus groups (*n*=20) and usability tests (*n*=15) reported using the internet everyday. In terms of digital literacy, caregivers generally reported more confidence with the use of digital technologies (*M*=8.56, *SD*=1.26) than healthcare professionals (*M*=7.83, *SD*=1.83).

### 3.2 Usability Evaluation

The mean SUS score for both caregivers and healthcare professionals was 74.3 (*SD*=18.0), which represented good usability of iSupport-Malaysia platform (Bangor et al., 2008). However, caregivers generally perceived its usability more favourably (*M*=76.1, *SD*=20.5) than healthcare professionals (*M*=71.7, *SD*=14.8).

In terms of task performance (see details in **Table 2**), participants successfully completed most tasks without assistance (>50%) on the iSupport-Malaysia website, though 33.3% did not complete the initial sign-up process due to problems with password requirements, forgotten email and password details, and technical problems on the platform. Nearly half of the participants did not understand how to ‘search’ for a specific lesson matching their needs (46.7%), as they either could not locate the ‘Search’ button, or tended to insert long phrases instead of lesson keywords in the search box. Participants also reported difficulties in locating the link to view or select a specific lesson, and needed prompts from the facilitator (46.7%). Nonetheless, once landed on the designated lesson page, most were able to follow through and complete the designated lesson independently (≥80.0%). Specifically, they were able to locate and watch lesson videos (80.0%), complete quizes (86.7%), and download lesson handouts (80.0%) without any assistance.

**Table 2.** Task completion rates, participants’ perception of task difficulties (1 = very difficult; 5 = very easy), and type of errors observed while performing the ‘Think Aloud’ tasks during usability tests (*n*=15).

| Task | Completed without assistance, $n$ (%) | Completed with assistance, $n$ (%) | Not completed, $n$ (%) | Difficulty Rating, $M$ ($SD$) | Type of Errors Observed |
| --- | --- | --- | --- | --- | --- |
| **1. Sign up** | 6 (40.0) | 4 (26.7) | 5 (33.3) | 3.60<br>(0.99) | • Password creation not complying to requirements<br>• Had own OpenLearning account but could not remember their email/password<br>• OpenLearning platform technical problems |
| **2. Find a specific lesson** | 8 (53.3) | 7 (46.7) | 0 (0) | 3.50<br>(1.18) | • Could not locate the link for lesson access<br>• Clicked on the 'lesson overview' diagram instead of lesson link |
| **3. Watch lesson video** | 12 (80.0) | 3 (20.0) | 0 (0) | 3.93<br>(1.03) | • Could not locate the video (mistaken video as an image)<br>• Could not find the 'full screen' button |
| **4. Complete lesson quiz** | 13 (86.7) | 2 (13.3) | 0 (0) | 3.93<br>(0.70) | • Did not click 'Submit Answer' because could not see the button<br>• Did not know could select more than one answer<br>• Difficulty reading quiz feedback (wordy, small font) |
| **5. Download lesson handout** | 12 (80.0) | 3 (20.0) | 0 (0) | 4.27<br>(0.88) | • Could not locate the 'download' link (small font, lack of contrast) |
| **6. Use "Search"** | 8 (53.3) | 4 (26.7) | 3 (20.0) | 3.73<br>(1.28) | • Could not locate the "Search" button<br>• Typed in English instead of Malay language<br>• Inserted long phrases to 'Search' rather than keywords |
| **7. Progress checking** | 9 (60.0) | 6 (40.0) | 0 (0) | 3.73<br>(1.16) | • Could not locate the progress bar<br>• Did not notice the small pie chart on top of each module |

All usability tasks were rated in the range of ‘moderate’ to ‘easy’ (rating range: 3.50–4.27). The task perceived as the easiest was downloading a lesson handout (mean rating: 4.27±0.88), followed by completing lesson quizzes (mean rating: 3.93±0.70) and watching lesson videos (mean rating: 3.93±1.03). By contrast, locating a specific lesson was perceived as the most challenging (mean rating: 3.50±1.18).

### 3.3. Qualitative Findings (Participants’ Insights)

To explore how users perceived the experience and educational content of iSupport-Malaysia, qualitative feedback from caregivers and healthcare professionals was thematically analysed. Since caregivers’ and healthcare professionals’ feedback from both focus groups and usability tests were similar, these were merged and presented under four overarching themes together: (i) teaching and learning experience; (ii) navigation of virtual learning; (iii) appropriateness of content; and (iv) program credibility and usefulness. The themes and subthemes were mapped in Figure 3. Within each theme, shared and divergent perspectives emerged between caregivers and healthcare professionals.

**Figure 3.**
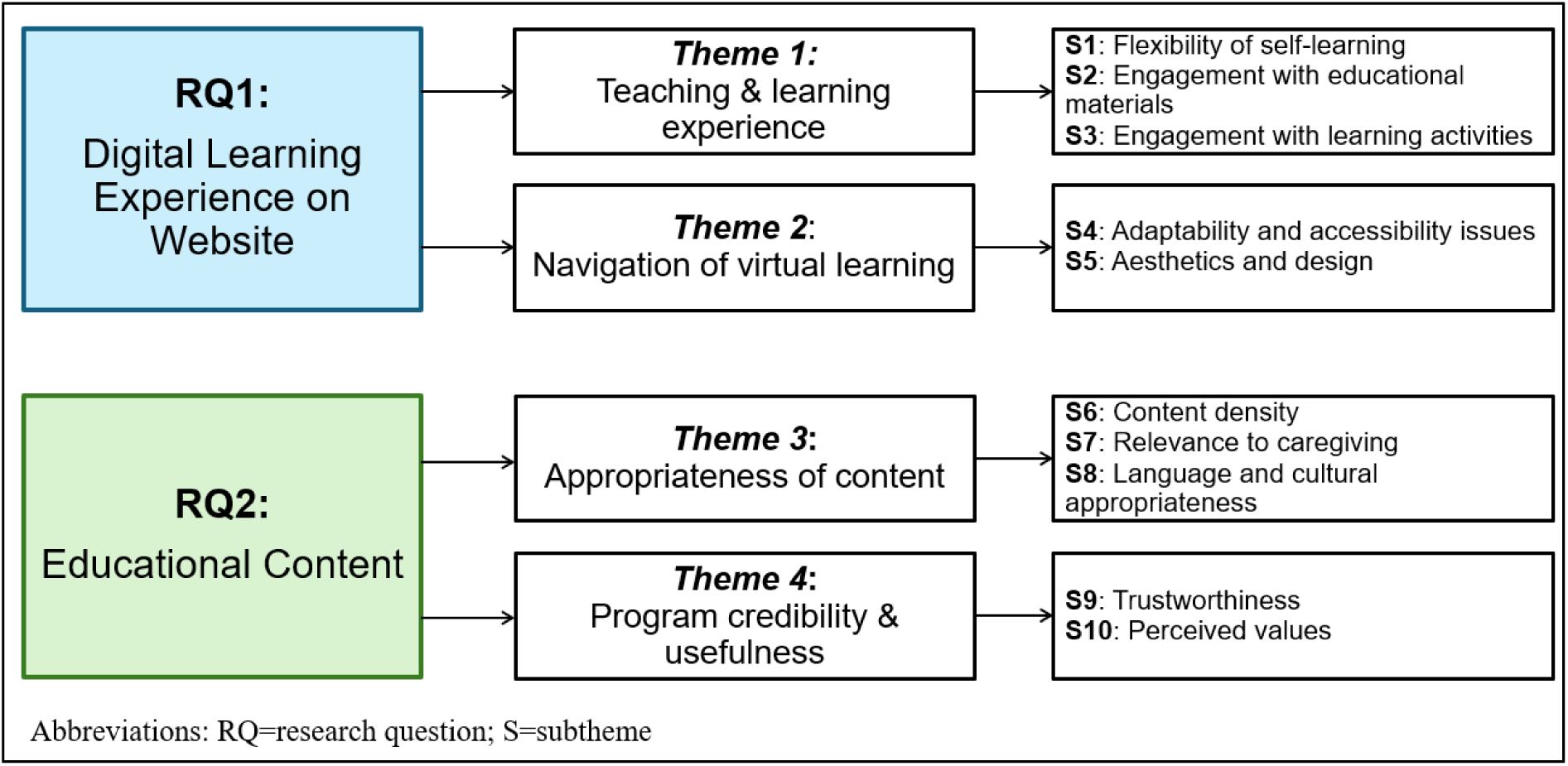
Themes and subthemes answering each research question.

### Theme 1: Teaching and learning experience

#### Flexibility of self-learning

Caregivers generally appreciated the flexibility of iSupport’s self-paced design, noting that the ability to choose lessons based on immediate needs was especially helpful given their busy schedules and time constraints.

> “*This is very good, flexible… Say, I’m very stressed and burnt out. So, maybe I can study module three first*.” [CG-UT05]

However, healthcare professionals raised concerns that skipping modules might result in caregivers missing certain essential foundational knowledge.

> *“… we need to re-emphasise about the importance of routine again and again for different modules… because if they don’t start (the modules) from the beginning, then it’s hard for them to capture the fundamentals…”* [HP-UT06]

#### Engagement with educational materials

Educational materials encompassed lesson videos and handouts, which were a one-way delivery of information without further user interaction. Notably, the use of animations in lesson videos drew mixed responses. Healthcare professionals viewed cartoon-style videos as engaging, whereas many caregivers found them distracting and less relatable. Some caregivers also described the videos as overly lengthy.

> *“I find it* (the videos) *quite relaxing and entertaining… it’s quite helpful, because nowadays, even the elderly are watching TikTok… They love this type of animation…”* [HP-FG09]
>
> *“I find the cartoons a bit distracting, because sometimes they* (the cartoon characters) *will be doing the same thing… it was not helpful because we are talking about something serious.”* [CG-FG01]

However, both caregivers and healthcare professionals agreed that the downloadable handouts were practical tools that distilled lessons into bite-sized tips and accessible takeaways.

> *“The handout is good… we can print out, give the patient and ask them to read while we’re reviewing clinical notes…”* [HP-FG01]
>
> *“The attachments that can be downloaded, it’s good. It serves as a guide for us.”* [CG-FG09]

#### Engagement with learning activities

Quizzes and reflection exercises were inserted as learning activities in most lessons to reinforce learning. First, quizzes received negative feedback from both caregivers and healthcare professionals. Participants felt that the quiz format offered little reassurance, as the right/wrong scoring risked invalidating caregivers’ efforts and their daily struggles. Some of the feedback was also perceived as lacking practical guidance.

> *“*… *maybe just spin the (quiz) feedback in a slightly more positive way… even though they* (the caregivers) *didn’t pick all the correct answers, the feedback can say they didn’t get it totally wrong either…”* [HP-FG07]
>
> *“*(For quizzes) *Scoring would not be necessary… I prefer you give me more options of other possible ideas and solutions to help me…”* [CG-UT05]

Reflection exercises were appraised more positively. For exercises with ‘comment’ features, caregivers especially appreciated the opportunities for peer sharing as a way of learning from one another.

> *“There was like the opportunity to share, (for example) what do you feed your loved ones?… I really like that… it’s a good opportunity for caregivers to exchange tips…”* [CG-FG03]

Nonetheless, exercises presented in a PDF worksheet format were perceived as less useful in the absence of facilitator interaction.

> *“I think some of the worksheets may require the caregivers to do it together with the professional… without any professionals available, it might be difficult for caregivers to even attend to it… they probably would just give up.”* [HP-FG03]

### Theme 2: Navigation of virtual learning

#### Adaptability and accessibility issues

The user-friendliness of the iSupport website varied across devices. On desktop, laptop and tablet, the website was easy and intuitive to navigate. However, on mobile browsers, participants encountered navigational difficulties, citing issues such as small fonts, inconsistent layouts, and overlapping icons.

> “*I actually quite like OpenLearning. It’s quite easy to navigate and very easy to find where you’re going…”* [HP-FG06]
>
> *“On Notebook, everything is simple, very easy to navigate… I give up (trying to browse the website) using mobile phone, because I felt like, a bit hard to navigate…”* [CG-FG05]

Beyond technical factors, participants recurrently highlighted age, digital literacy and educational backgrounds as additional barriers. In particular, older adults and those unfamiliar with online educational platforms might face greater challenges when using the program.

> “…*there are people who are not very good in handling all these yet, especially those in their 60s or 70s… If they are younger generation, maybe around 40, 50, they don’t need much technical help*…” [HP-FG06]
>
> *“OpenLearning is meant for people who are in higher education and learning… But, your target audience are basically elderly people like me and my wife… it could be difficult for us to learn…”* [CG-UT02]

#### Aesthetics and design

Website aesthetics, particularly the colour scheme and font, were viewed as important factors affecting usability experience among the middle- and older-age groups. The jade green background and white palette were originally used as the theme colour for its calming ambience. Nonetheless, this theme colour was described as less striking, less attractive, and hard to read. Inconsistent fonts also increased the difficulties for identification of key information.

> “*The problem is, when entering iSupport (website), the fonts, colours, sizes are different… Then I get a bit lost… It’s not standardised… It’s hard to find where to click* …” [CG-FG07]
>
> *“As someone who’s caring for a patient with dementia, I would be in the 50s or 60s. I might have issues with my eyesight, and I think the colour contrast and inconsistent font here can be a bit challenging for reading…”* [CG-UT08]

### Theme 3: Appropriateness of content

#### Content density

Opinions were divided regarding the amount of content in iSupport. While an equal split of healthcare professionals and caregivers considered the content adequate and comprehensive, others described it as overwhelming and wordy. It was noted that caregivers’ demanding schedules limited their capacity to absorb lengthy content.

> *“(The content) is a bit too long and chunky. People don’t have that kind of attention span… a person taking care of dementia (person) is quite desperate. It’s not like, I can take time to learn and understand (the lessons).”* [HP-FG04]
>
> “*I prefer… a shorter content… especially if you’re also working and, you know, doing a lot of stuff. This (the lessons) is kind of too long*.” [CG-FG06]

#### Relevance to caregiving

Caregivers differed in their perceptions of the practical relevance of the content. A small fraction of them found the content helpful, while others felt it did not always reflect the variability of real-life caregiving, as “*each dementia patient is different*” [CG-UT02].

> “*The example or the solution… I don’t think it’s actually helpful for me, because, unfortunately, my mum is a little bit stubborn…”* [CG-FG02]

To improve relevance, healthcare professionals suggested including additional practical topics such as assistive equipment, feeding positions, de-escalation strategies, and managing comorbidities. They also recommended inserting links to external resources and professional services.

> *“About the resources… for example, if the patient becomes too aggressive, or when the caregiver becomes too depressed, is there any external link that can direct them to other director[ies] to get help?”* [HP-FG10]

#### Language and cultural appropriateness

The Malay language used in videos and text materials was generally easy to understand. However, certain lessons reportedly contained formal language and medical jargons, which might demotivate users from continual usage.

> *“… some of the sentences… the way it was phrased was a bit awkward… it sounded like someone just directly translate for me… it’s not very smooth.”* [CG-FG05]
>
> “*Technical jargons, they may not be able to understand… (but) we can include the definition. Maybe instead of “vaskular”, we can use “saluran darah”* (blood vessels)… *or “infeksi” (become) “jangkitan”*. *So, it will be easier to understand*.” [HP-FG05]

Examples within the lessons were considered relevant to the local, multiracial and multicultural context. Nonetheless, one participant pointed out implicit gender bias in the content, as the examples appeared to imply caregiving roles as woman-oriented. Another participant called for broader inclusion of gender perspectives in caregiving,

> “*The thing that bothers me a lot is we keep giving examples as if a woman has to quit her job to be a full-time caregiver… We need to get people used to having men also be part of the dementia journey*.” [HP-FG04]
>
> “… *The challenges between a male caregiver and a female caregiver are different… there are certain things that a male caregiver can do that the female caregiver cannot do*…” [CG-UT02]

### Theme 4: Program credibility and usefulness

#### Trustworthiness

Generally, participants perceived iSupport to be a credible resource to learn about dementia caregiving, as the content was developed by renowned local universities. Nonetheless, participants suggested enhancing program trustworthiness by including caregivers’ testimonials and facilitator profiles.

> *“Trustworthiness… first thing that I saw is the university logos up there. That is a plus point already. And I know about OpenLearning, it’s trustworthy… But why is there no face (photos) for the team? Photos will look more professional…”* [HP-UT02]
>
> *“(Insert) testimonials or images. Because graphics speak louder than words… Team is also very important because who’s behind the project? Are you credible? Are you actually a voice of authority in the space of dementia? “* [CG-UT09]

#### Perceived values

Both healthcare professionals and caregivers endorsed iSupport as a valuable educational platform. Healthcare professionals saw it as a practical complement to clinical care, where limited consultation time often restricted opportunities for caregiver education.

> *”…like emotional management, self-care of the carer themselves, and other available resources… we may not be able to cover all of these during clinic consultation session. So, I think this is a good platform to educate and also support the caregiver*.” [HP-FG05]

For caregivers, iSupport was seen as having the potential to increase awareness, promote knowledge sharing, and reduce stigma around dementia. Nevertheless, the current program was perceived as more suitable for early-stage dementia, and participants called for content enhancement that is tailored to mid- and late-stage dementia.

> *“… if somebody recently received a diagnosis, or like someone suspects that their parents may have dementia, I think it’s a good platform to recommend to them, because all the information is in one place*.” [CG-FG03]
>
> *“… the way the modules are structured, it is assuming that the caregiver, as well as the dementia patient is at the beginning stage of their journey together… while a lot of us here are (taking care of) either the mid or end-stage dementia… I’d suggest… make your current modules the beginner content… And then you can have the intermediate content or, you know, advanced level content (to cater for mid- and late-stage dementia).”* [CG-FG08]

## 4. Discussion

This study presented the cultural adaptation and evaluation process for the preliminary iSupport-Malaysia online psychoeducation program. By adapting Barrera and Castro’s (2006) cultural adaptation framework (Barrera & Castro, 2006), the generic iSupport was localised through careful translation and content modification. While preserving the core concepts of each lesson, changes were made in terms of names, terminology, and customs relevant to local practice. The text-heavy format of iSupport was transformed into a more engaging multimedia format, designed to be accessible on an e-learning platform. Subsequently, this preliminary version of the iSupport-Malaysia e-learning platform was evaluated by caregivers and healthcare professionals to gather in-depth user insights.

Overall, our preliminary iSupport-Malaysia program was considered usable by caregivers and healthcare professionals, as demonstrated by a good (but not excellent) mean usability score of 74.3. More than half of the participants were able to complete most usability tasks without assistance from the facilitator. Qualitative findings further highlighted its potential in terms of teaching and learning, as well as content appropriateness and usefulness. Interestingly, differences in viewpoints were observed between the stakeholders, especially for the subthemes on self-learning flexibility, content density and practical relevance. This difference could be attributed to caregivers emphasising emotional and practical challenges driven by their first-hand caregiving experiences, whereas healthcare professionals focused on evidence-based practices and intervention recommendations based on their clinical expertise (Morgan et al., 2023).

One desirable feature of iSupport-Malaysia was the program’s self-paced flexibility, which was generally appreciated by caregivers in this study as it allowed them to decide when and how to engage with their learning amid demanding daily responsibilities. However, healthcare professionals cautioned that this level of unguided flexibility might inadvertently overwhelm users, leading users to miss out on certain foundational knowledge, thus recommending the modules to be learned sequentially. These findings have important implications on program engagement. Yardley et al. (2015) (Yardley et al., 2015) revealed that unrestricted navigational freedom could reduce adherence, whereas structured ‘tunnelling’ approaches that guide users through essential content might improve engagement. These insights underscore the importance of an initial needs assessment conducted by the program referrer or a designated program coordinator. For example, the care recipient’s stage of dementia and the caregivers’ current caregiving challenges shall be assessed before providing tailored module recommendations to better motivate users in a self-learning online environment.

Several iSupport adaptation studies, which presented the iSupport content in text-based format, highlighted caregivers’ preference to learn about dementia caregiving in engaging formats like videos and audios (Baruah et al., 2020; Molinari-Ulate et al., 2023; Xiao et al., 2022). Accordingly, our iSupport-Malaysia employed animation videos in simple, conversational language to enhance accessibility, particularly for caregivers with language barriers (Turk et al., 2019). Nonetheless, our findings caution that lengthy videos, with an excessive use of animations, may distract learners from capturing the key points. Prior studies recommend keeping videos under six minutes, using simpler visual elements, and presenting one key idea per video (Brame, 2016). In addition, there was mixed feedback on the quiz format. While the quiz scenarios were relatable, the ‘correct/incorrect’ scoring was seen as insensitive and invalidating. Similar concerns have been raised in other iSupport adaptations (Masterson-Algar et al., 2022; Morgan et al., 2023), where quizzing exercises were perceived as judgemental, since the ‘correct’ answers may not work for every person with dementia. Therefore, our program refinement should consider reframing feedback in a more supportive tone, offering more practical suggestions, and incorporating more peer-sharing features to promote peer support and learning (Wen et al., 2022).

Next, usability and accessibility were essential factors influencing the adoption of web programs. Our study found a mean System Usability Scale (SUS) score of 74.3, indicating good usability (Bangor et al., 2008; Brooke, 1996). However, it was notably lower than those reported in iSupport-Brazil’s (86.5) and iSupport-Portugal’s (89.5) usability studies (Ottaviani et al., 2022; Teles et al., 2021). This difference may be due to the integration of multimedia elements in our e-learning platform, in contrast to the text-and-slide formats utilised in the adaptations from Brazil and Portugal. Our usability testing uncovered two major challenges: the sign-up process and search function, which had non-completion rates of 33.3% and 20% respectively. These findings align with literature showing that complex registration procedures and unintuitive navigation can discourage engagement, leading to discontinuation (Hassan, 2020). To mitigate these issues, we propose inserting step-by-step tutorials for the sign-up process and essential navigation features within the platform. A dedicated program coordinator can also be appointed to assist users in technical queries. Furthermore, enhancements to the aesthetics of the platform, such as use of clear clickable targets, consistent content presentation, and readable fonts with strong contrast should also be considered to increase intuitiveness and ease of navigation (Hattink et al., 2016).

Our study also highlighted content relevance to be another key factor influencing program engagement. Caregivers and healthcare professionals alike appreciated the comprehensiveness of the modules and the culturally sensitive case scenarios presented in simple local language. However, certain lessons were viewed as overwhelming or not directly applicable to the caregivers’ current situations. Additionally, the lack of gender-specific scenarios diminished the relevance for some, particularly since earlier studies have shown that male caregivers tend to favour problem-focused approaches, in contrast to the emotion-focused strategies preferred by female caregivers (Wen et al., 2022). Meanwhile, healthcare professionals suggested expanding the content to include assistive equipment, de-escalation techniques, and other information that caregivers ‘should’ know. These divergent priorities – caregivers seeking immediate, practical solutions while professionals emphasising comprehensiveness – illustrate the need for tailored support. Implementing dialogue functions, such as AI chatbots or live consultations, could help meet these differing needs without overwhelming users (Islas-Cota et al., 2022). In fact, studies on dementia caregivers have expressed desires for professional or peer interactions to reduce isolation and provide reassurance (Molinari-Ulate et al., 2023; Xiao et al., 2022).

Our findings suggest iSupport-Malaysia has the potential to enhance existing caregiver support structures. Caregivers appreciated it as a structured, accessible, and flexible alternative to fragmented information online; while professionals view it as a valuable addition to their clinical practices, facilitating support and knowledge sharing between patient visits. This dual benefits position iSupport as a cost-effective, scalable resource, particularly in regions with limited access to dementia care and psychosocial support (Loh et al., 2024). To maximise reach, however, our caregivers and caregivers from other iSupport adaptations have recommended future research to expand the content to cover moderate and late-stage dementia (Baruah et al., 2021; Teles et al., 2021). Additionally, program credibility also relied on visible accreditation from reputable institutions, expert involvement, and caregiver testimonials, consistent with findings that social proof and institutional backing significantly bolster trust in eHealth interventions (Sbaffi & Rowley, 2017). Consequently, promoting iSupport through healthcare settings such as primary care clinics, memory clinics, and hospitals may encourage uptake more effectively than mass advertising.

To the best of our knowledge, this study is the first among the WHO’s iSupport adaptation studies to evaluate the user perception of a multimedia, e-learning version of iSupport. This study underscores the value of usability testing and mixed methods evaluation during eHealth intervention development. Observing users in real time allowed us to identify navigation challenges, technical problems and user confusion – issues that might not be captured from focus groups or interviews alone. Engaging stakeholders early is crucial to ensure the intervention is motivating, enjoyable, informative, and has the potential to enhance wellbeing (Lee et al., 2022; Yardley et al., 2015). Insights gathered from this study could inform the adaptation refinement of our current eHealth intervention, where key design principles such as the use of shorter, focused videos; enhancement of aesthetic features; simplified navigation without complex logins; and person-centred tailoring of recommendations with emotionally sensitive framing would be considered when improving the current iSupport-Malaysia platform.

Additionally, this study provides new insights into how culturally adapted digital psychoeducation can be optimised for Malaysian caregivers. Unlike existing local resources such as the Demensia-KITA app, which primarily delivers text-based content and English-language videos with Malay subtitles (Rashid et al., 2024), iSupport-Malaysia transforms caregiver training into fully localised video-based lessons in the national language, interactive learning activities, and incorporates peer-sharing features to foster social connectedness. These design elements were viewed as highly valuable, though stakeholders also emphasised the need to integrate professional support to enhance user trust (Hassan, 2020). The findings collectively demonstrate that, with further refinement, iSupport-Malaysia holds strong potential to be scaled as a national resource, directly supporting Thrust 2 of the National Dementia Action Plan, which prioritises accessible training and support for family caregivers in Malaysia (Ministry of Health Malaysia, 2024).

## Limitations and Future Recommendations

This study has several limitations. Firstly, while our sample size was adequate for exploratory purposes, caution should be taken when generalising the findings across Malaysia’s heterogeneous caregiver population. Although the sample was fairly diverse, our participants were still primarily urban (Selangor and Kuala Lumpur regions), with only two participants lacking college or university education. Given the link between education and internet use (Ottaviani et al., 2022), the challenges of rural or low-literacy caregivers may be under-represented. Besides, most participants were middle-aged individuals who might be more familiar with technology. Hassan (2020) (Hassan, 2020) emphasised the need to involve older caregivers in the design process, since technology solutions perceived as user-friendly for younger users might not be user-friendly for older users. In fact, the older participants from our usability testing reported reservations in using iSupport, citing issues with navigational complexity and limited mobile accessibility. Future studies should include a broader demographic spectrum, especially older adults and those from resource-constrained settings.

Additionally, this evaluation captured only a snapshot of user experiences at a single point in time. Hence, longitudinal studies are needed to assess sustained use and long-term outcomes. Further, a pilot randomised controlled trial is currently underway to evaluate the feasibility, acceptability, and preliminary effectiveness of the refined iSupport-Malaysia program (Loh et al., 2025). This will provide deeper insights into its potential for improving caregiver outcomes nationwide.

## Conclusion

This study is the first to evaluate the cultural adaptation and development process of a multimedia, e-learning adaptation of the WHO’s iSupport program. We offer novel insights into how local stakeholders perceived the culturally adapted iSupport intervention delivered via an engaging, interactive digital platform. By combining focus group discussions and usability tests from both caregivers and healthcare professionals, we identified several key strengths, including flexibility, cultural relevance, and potential to complement existing care; alongside areas for refinement in navigation, lesson delivery, and personalised support. The findings underscore the importance of grounding eHealth interventions in a culturally sensitive manner and user-centred design, ensuring they are both practical, relevant and trustworthy. With further development and refinement, iSupport-Malaysia holds promise as a scalable, accessible, and cost-effective resource that can strengthen dementia care support systems and contribute towards national priorities for caregiver wellbeing.

## Supporting information

Supplemental File 1: COREQ Checklist

Supplemental File 2: Focus Group Guide

## Acknowledgments

We would like to thank Aaron Koay Chee Cheng from Monash University Malaysia for proofreaeding the manuscript; and Khailesh Bushan a/l Balakrishnan from Universiti Sains Malaysia for his time and assistance in co-facilitating the focus groups. We also would like to thank all participants of this study for reviewing our iSupport-Malaysia platform and offering their unique insights. Additionally, we would like to extend our acknowledgement to Izrul Fizal, Katty Lou Jia Qi, and Aisyah binti Zulkipli for their contributions in producing the voiceovers and high-quality videos for iSupport-Malaysia; and OpenLearning technical support team for assisting in the development of the iSupport-Malaysia platform.

## Use of generative AI statement

ChatGPT (version 5.1) was used solely to refine wording and enhance language clarity during manuscript preparation. No generative AI tools were used to develop or write research questions, methodology, data analysis, and interpretation of findings. All content was reviewed and approved by the authors, who take full responsibility for the final manuscript.

## Author Contributions

WLL and KJL designed the methodological framework and data analysis plan. KJL completed data collection and data analysis with the support from WLL, ALON, FFLC and ER. All team members provided input for theme development and validated the results. KJL conceptualised and drafted the initial manuscript. WLL, FFLC, ALON and ER contributed to the content and revision of manuscript. All authors have read and approved the final version.

## Ethical Considerations

### Ethical Approval

This study was approved in full by the Medical Research Ethics Committee of University of Malaya (MRECID No.: 2023927-12912) and the Research Ethics Committee of Sunway University (REC ID: PGSUREC2023/083). The entire procedures were conducted in line with the Declaration of Helsinki principles.

### Consent to Participate

All participants who participated in the focus groups and usability tests were provided with information sheets to review, and all participants had provided written or electronic informed consent prior to participation. The online focus group sessions were audio- and video-recorded with participants’ consent. They were informed that they had the right to turn off their camera to respect their privacy, and they were encouraged to use short names during the session for anonymity. All recordings were downloaded and stored in the main researcher’s password-protected files.

### Consent for Publication

All participants provided informed consent for publishing aggregated, deidentified questionnaire results, as well as anonymised quotes from the focus groups and usability tests sessions.

## Data Availability Statement

All data produced in the present study are available upon reasonable request to the authors.

## Footnotes

a There was an initial sample of ten caregivers who provided electronic informed consent to participate in the focus groups (FGs). However, one dropped out prior to the focus group session citing insufficient digital literacy and difficulties navigating iSupport. The final sample of nine was subdivided into the two FGs following their availability. Group 1 consisted of four informal caregivers, whereas Group 2 consisted of three informal caregivers and two formal caregivers.

