## Supplemental File 2: Focus Group Guide for "Development and Evaluation of *iSupport-Malaysia*: A Multimedia Web-Based Psychoeducational Intervention for Dementia Caregivers"

### Supplemental File 3: Focus Group Guide

| Aspect | Stem Questions | Additional Prompts (when needed) |
|---|---|---|
| **1. Aesthetic ('look' and 'feel')** | Please tell us what you like or don't like in terms of our general website layout, layout of the quiz or reflection exercise, colour, font and font size, and diagrams used on the website. | Why do you like that…?<br><br>Why don't you like that? What would you suggest? |
| **2. Ease of Use** | 1. Which device did you use to access iSupport-Malaysia website?<br>Please 'raise' your hand if you used:<br>a) Mobile phone<br>b) Tablet (iPad, android tablets)<br>c) Computer desktop or laptop<br><br>2. Based on your user experience, please vote if you think most people can use iSupport:<br>a) Without any technical help<br>b) With minimal technical help<br>c) With more technical help<br><br>3. Please tell us why you think the iSupport website was very easy / difficult to use. | What was your experience on mobile phone / tablet?<br><br>What were some of the technical problems you encountered when using the website? |
| **3. Language clarity & cultural sensitivity** | 1. Could you tell us what you think about the language use in the texts, quizzes and videos of iSupport?<br><br>2. How easy or difficult was it to understand the Malay language used on the website?<br><br>3. Is there any part that you think is not appropriate for our Malaysian culture? Tell us more. | What can you suggest making the language plainer and easier to understand by most caregivers?<br><br>How would you suggest replacing the part that might be culturally inappropriate? |
| **4. Comprehen-siveness of content** | 1. We would like to hear your thoughts about:<br>- Whether the lessons have enough coverage<br>- Whether the lessons are well-organised<br>- Whether the videos and learning activities matched the learning objectives<br>- How interactive and engaging the lessons are | Is there any information that you find to be excessive or unnecessary on the website?<br><br>Is there any information lacking, and what's your recommendation for improvement? |

| | | |
|---|---|---|
| | - Whether the lessons are relevant and accurate *(this question was asked for healthcare professionals only)*<br><br>2. What do you think of these additional features on our website?<br>- Printable handouts/ exercises<br>- Comment boxes to share knowledge with others<br>- Lesson completion batches at the end of each lesson | Is there any website feature that you think is unnecessary and shall be removed from the website? |
| **5. User satisfaction and motivation to recommend the program** | <u>Caregivers' Group</u>:<br>1. Based on your user experience, will you continue or stop using iSupport-Malaysia in the next 2-3 months? Please vote:<br>A) I will continue using iSupport in the next 2-3 months<br>B) I will not continue using iSupport<br><br>2. Can you explain more what motivates you to/ stops you from using iSupport?<br><br>3. How likely would you recommend iSupport-Malaysia to other caregivers? Please vote:<br>A) Very likely<br>B) Likely<br>C) Unsure/ Unlikely<br><br><u>Healthcare Professionals' Group:</u><br>1. How likely would you recommend iSupport-Malaysia to your clients? Please vote:<br>A) Very likely<br>B) Likely<br>C) Unsure/ Unlikely<br><br>2. Can you please share with us what aspects of iSupport make you want to/ do not want to recommend iSupport to your clients? | Please share with us what aspects of iSupport make you feel unsure whether to recommend iSupport to other caregivers.<br><br>Please elaborate more on why you will (very) likely recommend iSupport to other caregivers. |
